# Comparison of Care Utilization and Medical Institutional Death among Older Adults by Home Care Facility Type: A Retrospective Cohort Study in Fukuoka, Japan

**DOI:** 10.1101/2020.06.21.20136085

**Authors:** Sung-A Kim, Akira Babazono, Aziz Jamal, Yunfei Li, Ning Liu

**Affiliations:** Department of Health Care Administration and Management, Graduate School of Medical Sciences, Kyushu University; Department of Health Care Administration and Management, Graduate, School of Medical Sciences, Kyushu University, Fukuoka, Japan; Health Administration Program, Faculty of Business and Management, Universiti Teknologi MARA, Selangor, Malaysia; Department of Preventive Medicine and Community Health, University of Occupational and Environmental Health, Kitakyushu, Japan

**Keywords:** Home care, integrated care, health care cost, long-term care, aging in place

## Abstract

**Objectives:** We compared the care services use and medical institutional deaths among older adults across four home care facility types.

**Design:** This was a retrospective cohort study.

**Setting:** We used administrative claims data from April 2014 to March 2017.

**Participants:** We included 18,347 residents of Fukuoka Prefecture, Japan, who received home care during the period, and aged ≥75 years with certified care needs of at least level 3. Participants were categorized based on home care facility use (i.e., general clinics, Home Care Support Clinics/Hospitals (HCSCs), enhanced HCSCs with beds, and enhanced HCSCs without beds).

**Primary and secondary outcome measures:** We used generalized linear regression models to estimate care utilization and the incidence of medical institutional death, as well as the potential influence of sex, age, care needs level, and Charlson comorbidity index as risk factors.

**Results:** The results of generalized linear models showed the inpatient days were 53.3, 67.4, 63.9, and 72.6 for users of enhanced HCSCs with beds, enhanced HCSCs without beds, HCSCs, and general clinics, respectively. Correspondingly, the numbers of home care days were 64.0, 51.6, 57.9, and 28.4. Our multivariable logistic regression model estimated medical institutional death rate among participants who died during the study period (*n* = 9919) was 2.32 times higher (*P*<0.001) for general clinic users than enhanced HCSCs with beds users (relative risks=1.69, *P*<0.001).

**Conclusions:** Participants who used enhanced HCSCs with beds had a relatively low inpatient utilization, medical institutional deaths, and a high utilization of home care and home-based end-of-life care. Findings suggest enhanced HCSCs with beds could reduce hospitalization days and medical institutional deaths. Our study warrants further investigations of home care as part of community-based integrated care.

**Trial registration:** This study was approved by the Kyushu University Institutional Review Board for Clinical Research (Approval No. 20209).

**Strengths and limitations of this study:**

1. This was a retrospective cohort study including data on 18,347 individuals.
2. This study was designed to suggest the kind of healthcare system that will be needed in the future in aging societies by examining the associations of the type of home care provision system with end-of-life care and place of death for older adults.
3. We calculated the number of years that participants lived during the study period and estimated the annual utilization rates per person-year of observation.
4. This study was conducted using data only on residents of Fukuoka Prefecture in Japan, which limits the generalizability of our findings
5. There were no clinical data for individual participants because this study focused on the types of healthcare facilities that provide home care.

## INTRODUCTION

In 2012, approximately 800 million adults—11% of the global population—were aged 65 years or older.[1] This age group is projected to reach 1.4 billion people by 2030 and to exceed 2 billion around 2050.[2] Japan is presently the world’s most aged society, with adults aged 65 years and older accounting for 22.6% of the country’s population in 2010, and this percentage is expected to surpass 30% in 2025.[3] Notably, the post-World War II baby-boom generation in Japan will reach the age of 75 years in 2025, imposing a heavy financial burden on the nation’s social security system.[4, 5]

### The present of care for the older adults in Japan

Japan’s health insurance system categorizes older adults aged 65–74 years as “early-stage elderly” and those aged 75 years and older as “latter-stage elderly,” and these age groups’ out-of-pocket copayment rates are set at 20% and 10%, respectively.[6] The average annual medical expenditure in 2018 was 553,000 Japanese yen (US$5,247) for early-stage elderly patients and 910,000 Japanese yen (US$8,634 and this is 1.6 times higher) for latter-stage elderly patients.[7] In contrast, the average annual long-term care (LTC) expenditure was 50,000 Japanese yen (US$474) for early-stage elderly patients and 480,000 yen (US$4,554 and this is over 10 times higher) for latter-stage elderly patients.[7] Latter-stage elderly patients tend to be relatively frail and to have multiple chronic conditions that require the use of both medical and LTC services.[8, 9] To efficiently provide integrated care for these individuals, Japan has implemented the Community-based Integrated Care System (CICS), with the aim of moving away from the conventional hospital-centered healthcare delivery system toward a system that is focused on patients’ residences and local facilities.[10] The CICS comprehensively provides medical care that provided at medical facilities, home care that provided at patient’s own house or nursing home by medical professional and LTC services such as day care at LTC facilities or home visiting by care workers in addition to preventive care and daily living support. These services enable older adults to age in place until the end of life, even when they become increasingly care-dependent.[11] It is necessary to ensure the availability of 24-hour, 365-day care services to monitor and manage any sudden changes in these older adults’ health status.

### Why is home-based end-of-life care necessary?

The percentage of deaths occurring at home in Japan exceeded 80% in 1951, with only 9% of deaths occurring at medical institutions such as hospitals.[12] This trend began to reverse in 1976, and 75.8% of deaths occurred in medical institutions in 2016,[12] despite approximately 70% of people reporting that they would prefer to spend the end of their lives at home rather than in a medical institution.[13] If the current trends continue, almost half a million people in Japan will be unable to receive end-of-life care at a medical institution in 2030, even if the number of home deaths increases by a factor of 1.5.[14] To resolve this issue, in 2006, Japan introduced Home Care-Support Clinics/Hospitals (HCSCs), which provide 24-hour home care and home-visit nursing care.[15] Furthermore, in 2012, Japan established “enhanced HCSCs,” which fulfill more stringent criteria such as having three or more full-time doctors on staff and having handled at least five cases of emergency home care treatment and at least two cases of end-of-life care within the past year.[15] HCSCs that qualify for this “enhanced” designation receive higher reimbursements compared with conventional HCSCs.[15] At present, general clinics, HCSCs, and enhanced HCSCs are authorized to provide insurance-covered home care. Enhanced HCSCs are further categorized into those with beds and those without beds,[16] yielding a current total of four types of home care facilities available in Japan.

### What affects the end-of-life care of older adults?

The percentage of adults aged 75 years and older in Japan is expected to reach 18.1% in 2025,[3] and optimizing community-based care systems may help to provide solutions for problems faced by aging populations in Japan and throughout the world. To improve the circumstances allowing older adults to continue living at home, it is necessary to first ascertain how different types of facilities in the current home care delivery system influence the use of medical and LTC services. Previous studies have shown that strengthening home care services has contributed to reducing patient hospitalization.[17-24] However, it remains unclear whether specific measures to strengthen the home care delivery system, such as the introduction of HCSCs and enhanced HCSCs, have affected where older adults receive end-of-life care or their utilization of various care services.

This study examined the influence of the home care delivery system on end-of-life care in adults aged 75 years and older. We comparatively examined home-based end-of-life care utilization, deaths in medical institutions, and the use of medical and LTC services among older adults who received home care services from four different types of facilities (enhanced HCSCs with beds, enhanced HCSCs without beds, HCSCs, and general clinics). In this study, general clinics refer to facilities other than HCSCs that provide home medical care services. Unlike HCSCs, these facilities are not eligible for insurance claims reimbursement due to establishment status.

## METHODS

### Database

The study was conducted using data from a medical claims database and an LTC insurance claims database provided by the Fukuoka Prefecture Association of Latter-stage Elderly Healthcare. Medical claims included information on patient characteristics, medical treatments, disease diagnoses, and medical expenditures for all individuals who received insurance-covered care.[25] LTC insurance refers to the public insurance for older adults aged ≥65 years and adults aged ≥40 years with specific diseases. These claims include information on LTC service utilization and the corresponding expenditures for all individuals with certified care needs. Under the LTC insurance system, care needs are categorized into seven levels (support needs levels 1–2 and care needs levels 1–5), with increasing levels signifying higher degrees of dependence.[25]

The administrative claims data were de-identified by constructing specific databases using a work station with no connection to any networks.

### Study Design

This retrospective cohort study used data from April 2014 to March 2017. The study participants were Fukuoka Prefecture residents aged 75 years and older with certified care needs of level 3 or higher in April 2014 who received home care services between April and June 2014. Residents who migrated to other Prefectures between April 2014 and March 2017 were excluded. The participants were divided into four groups according to the facility type providing them with home care services: Group A (enhanced HCSCs with beds), Group B (enhanced HCSCs without beds), Group C (HCSCs), and Group D (general clinics).

We conducted pooled cross-sectional study for participants who died during the 3-year study period to compare their home-based end-of-life care utilization and place of death across the four groups. The use of home-based end-of-life care was identified using claims records of additional fees specifically for these services. Place of death was categorized as medical institution for participants who were recorded as dying at a hospital or a clinic in the claims data. The medical institutional death rate was calculated as the percentage of all participants who died during the study period whose death occurred in a medical institution.

We also examined the number of days that participants received inpatient care, outpatient care, and home care across the four groups. Expenditures for inpatient care, outpatient care, home care, drug prescription and LTC services were also calculated for each group. Expenditures were converted from Japanese yen to US dollars using the 2017 purchasing power parity rate (US$1 = JP¥105.4).

Information was obtained on participant sex, age, care needs level, and Charlson comorbidity index (CCI) score as of April 2014.[26] Age was divided into four categories (75–79, 80–84, 85–89, and ≥90 years). The care needs levels included in our analysis were levels 3, 4, and 5 (with level 5 representing the highest level of dependence). CCI scores, which indicate the weighted number of concomitant diseases in an individual, were divided into three categories (0–2, 3–4, and ≥5).

### Patient and Public Involvement

We used administrative claims data and did not involve patients in this study.

### Statistical Analysis

The distributions of sex, age, care needs level, CCI, and death were examined across the four facility type groups. Additionally, inter-group differences in home-based end-of-life care utilization and institutional death among those who died during the study period were examined. One-way analysis of variance was used to compare these differences.

We constructed multivariable logistic regression models to evaluate the influence of home care facility type on home-based end-of-life care utilization and medical institutional death. In these models, the dependent variables were the use of home-based end-of-life care and death at a medical institution. The exposure of interest was the home care facility type, with Group A (enhanced HCSCs with beds) as the reference category. The covariates were sex, age, care needs level, and CCI. Odds ratios (ORs) and 95% confidence intervals were estimated.

Next, we calculated the mean annual days of inpatient care, outpatient care, and home care, as well as the mean annual expenditures for inpatient care, outpatient care, home care, drug prescription and LTC services for each facility type group. The inter-group differences were compared using analysis of variance. Each participant’s service utilization was calculated over the number of years he/she lived during the study period, and annual utilization rates per person-year of observation were estimated. This method allowed the inclusion of data from participants who died during the study period, which was useful because the study population comprised individuals with an elevated mortality risk because of advanced age and high care needs.

To evaluate the influence of home care facility type on the use of medical and LTC services, we constructed generalized linear models (GMLs). Here, the dependent variables were the numbers of days of inpatient care, outpatient care, and home care, as well as the expenditures for inpatient care, outpatient care, home care, drug prescription and LTC services. The exposure of interest was the home care facility type. The covariates were sex, age, care needs level, CCI, death, and the number of years the participant lived.

The dependent variables data were highly skewed and over-dispersed. Thus, analyzing these data using a conventional regression method might violate the data normality assumption. Furthermore, the data containing the number of days is commonly regarded as a ‘count’ variable, and the use of statistical techniques based on normal distribution might not be appropriate [27,28]. Many researchers have suggested the use of the generalized linear model (GLM) by assuming such data has a negative binomial or a Poisson distribution. [29,30,31]. In contrast, the use of GLM with a gamma distribution is recommended when analyzing data involving health care costs [32,33]. In our preliminary analyses, dependent variables containing the number of days were fitted in two separate models: GLM with a negative binomial distribution, and GLM with a Poisson distribution. Diagnostic statistics, however, identified GLM with a negative binomial distribution provides better estimates than the model with an assumed Poisson distribution. The results of the analysis presented in this study are therefore, based on the estimates of GLM with a negative binomial distribution with log-link function and robust standard errors, for the analyses involving the number of days. On the other hand, for the results of analyses involving care expenditure, the estimates of GLM with a gamma distribution with log-link and robust standard errors are presented.

The marginal means of the dependent variables were calculated to indicate the estimated values of numbers of care days and expenditures for the examined care services. These were calculated by substituting the mean of the estimates into GLMs with a negative binomial distribution for care days and a gamma distribution for care expenditure.

SQL Server 2014 was used to extract the data, and Stata, 14.2 was used for all analyses.

## RESULTS

The participants’ characteristics are summarized in table 1. The included participants were 18,347 Fukuoka Prefecture residents who used home care at any of the four facilities categorized in this study, between April and June 2014. Participants with a care of needs lower than 3 as of April 2014 and those who emigrated to other Prefectures during the follow-up period were excluded. The participants were comprising Group A with 2,509, Group B with 825, Group C with 6,218 and Group D with 8,795 participants who had both medical claims data and an LTC insurance claims data. We observed significant inter-group differences in sex (*P*=0.002), age (*P*<0.001), and CCI (*P*<0.008). However, there were no significant differences in care needs level (*P*=0.816) or death (*P*<0.669). Groups A and B tended to have higher CCI scores and older ages than did Groups C and D. During the 3-year study period, 54% of the participants died; Group A had the highest percentage of deaths (59.9%). Among the participants who died, there were significant inter-group differences in home-based end-of-life care utilization and in medical institutional death. Group A had the highest home-based end-of-life care utilization rate (57.4%) and the lowest rate of medical institutional death (25.6%). The home-based end-of-life care utilization rate in Group A, was followed (in order) by Groups B, C, and D, but this order was reversed for the rate of medical institutional death.

**Table 1.** Participant characteristics by home care facility type

|  | Total | Group A | Group B | Group C | Group D | *P* value |
| --- | --- | --- | --- | --- | --- | --- |
| Number of participants | 18,347 | 2,509 (13.7) | 825 (4.5) | 6,218 (33.9) | 8,795 (47.9) |  |
| Sex |  |  |  |  |  |  |
| Men (%) | 4,709 (25.7) | 645 (25.7) | 204 (24.7) | 1,473 (23.7) | 2,387 (27.1) | 0.002 |
| Women (%) | 13,638 (74.3) | 1,864 (74.3) | 621 (75.3) | 4,745 (76.3) | 6,408 (72.9) |  |
| Age |  |  |  |  |  |  |
| Mean [SD] | 87.5 [6.2] | 87.8 [6.1] | 88.1 [6.1] | 87.7 [6.1] | 87.1 [6.3] | <0.001 |
| 75–79 (%) | 2,051 (11.2) | 226 (9.0) | 70 (8.5) | 605 (9.7) | 1,150 (13.1) |  |
| 80–84 (%) | 4,035 (22.0) | 542 (21.6) | 167 (20.2) | 1,388 (22.3) | 1,938 (22.0) |  |
| 85–89 (%) | 5,394 (29.4) | 746 (29.7) | 258 (31.3) | 1,876 (30.2) | 2,514 (28.6) |  |
| ≥ 90 (%) | 6,867 (37.4) | 955 (39.7) | 330 (40.0) | 2,349 (37.8) | 3,193 (36.3) |  |
| Care needs levels |  |  |  |  |  |  |
| Level 3 (%) | 5,081 (27.7) | 582 (23.2) | 227 (27.5) | 1,739 (28.0) | 2,533 (28.8) | 0.816 |
| Level 4 (%) | 6,804 (37.1) | 882 (35.2) | 281 (34.1) | 2,341 (37.6) | 3,300 (37.5) |  |
| Level 5 (%) | 6,462 (35.2) | 1,045 (41.6) | 317 (38.4) | 2,138 (34.4) | 2,962 (33.7) |  |
| Charlson comorbidity index |  |  |  |  |  |  |
| 0–2 (%) | 4,115 (22.4) | 507 (20.2) | 144 (17.4) | 1,331 (21.4) | 2,133 (24.2) | 0.008 |
| 3–4 (%) | 6,629 (36.1) | 873 (34.8) | 295 (35.8) | 2,385 (38.4) | 3,076 (35.0) |  |
| ≥ 5 (%) | 7,603 (41.5) | 1,129 (45.0) | 386 (46.8) | 2,502 (40.2) | 3,586 (40.8) |  |
| Death |  |  |  |  |  |  |
| Yes (%) | 9,919 (54.1) | 1,502 (59.9) | 471 (57.1) | 3,271 (52.6) | 4,675 (53.2) | 0.699 |
| No (%) | 8,428 (45.9) | 1,007 (40.1) | 354 (42.9) | 2,947 (47.4) | 4,120 (46.8) |  |
| Number of deaths | 9,919 | 1,502 (15.1) | 471 (4.8) | 3,271 (33.0) | 4,675 (47.1) |  |
| Home-based end-of-life care |  |  |  |  |  |  |
| Yes (%) | 3,103 (31.3) | 862 (57.4) | 220 (46.7) | 1,285 (39.3) | 736 (15.7) | <0.001 |
| No (%) | 6,816 (68.7) | 640 (42.6) | 251 (53.3) | 1,986 (60.7) | 3,939 (84.3) |  |
| Medical Institutional death |  |  |  |  |  |  |
| Yes (%) | 3,633 (36.6) | 384 (25.6) | 137 (29.1) | 1,039 (31.8) | 2,073 (44.3) | <0.001 |
| No (%) | 6,286 (63.4) | 1,118 (74.4) | 334 (70.9) | 2,232 (68.2) | 2,602 (55.7) |  |

NOTES: Group A comprised users of enhanced Home Care Support Clinics/Hospitals (HCSCs) with beds, Group B comprised users of enhanced HCSCs without beds, Group C comprised users of HCSCs, and Group D comprised users of general clinics. "Number of deaths" refers to participants who died during the study period.

Table 2 shows the multivariate logistic regression analysis results for the associations of home care facility type with home-based end-of-life care utilization and medical institutional death among the participants who died during the study period. Relative to Group A, Group D had the lowest odds of using home-based end-of-life care (OR=0.13; *P*<0.001) and the highest odds of institutional death (OR=2.32; *P*<0.001).

**Table 2.** Associations of home care facility type with home-based end-of-life care utilization and medical institutional death

|  | Home-based end-of-life care |  | Medical Institutional death |  |
| --- | --- | --- | --- | --- |
|  | Odds ratio (95% CI) | P value | Odds ratio (95% CI) | P value |
| Home care facility type |  |  |  |  |
| Group A | Reference |  |  |  |
| Group B | 0.66 (0.53–0.82) | <0.001 | 1.17 (0.93–1.47) | 0.19 |
| Group C | 0.47 (0.41–0.54) | <0.001 | 1.35 (1.18–1.56) | <0.001 |
| Group D | 0.13 (0.11–0.15) | <0.001 | 2.32 (2.03–2.65) | <0.001 |
| Sex |  |  |  |  |
| Men | Reference |  |  |  |
| Women | 1.36 (1.22–1.51) | <0.001 | 0.75 (0.68–0.82) | <0.001 |
| Age |  |  |  |  |
| 75–79 (%) | Reference |  |  |  |
| 80–84 (%) | 1.11 (0.89–1.38) | 0.34 | 1.05 (0.88–1.25) | 0.58 |
| 85–89 (%) | 1.34 (1.09–1.65) | <0.001 | 0.95 (0.81–1.13) | 0.58 |
| ≥90 (%) | 1.96 (1.61–2.39) | <0.001 | 0.74 (0.63–0.87) | <0.001 |
| Care needs levels |  |  |  |  |
| Level 3 (%) | Reference |  |  |  |
| Level 4 (%) | 1.31 (1.15–1.49) | <0.001 | 0.88 (0.79–0.98) | 0.02 |
| Level 5 (%) | 1.94 (1.71–2.20) | <0.001 | 0.71 (0.63–0.79) | <0.001 |
| Charlson comorbidity index |  |  |  |  |
| 0–2 (%) | Reference |  |  |  |
| 3–4 (%) | 0.74 (0.65–0.84) | <0.001 | 1.25 (1.10–1.41) | <0.001 |
| ≥5 (%) | 0.57 (0.50–0.64) | <0.001 | 1.51 (1.35–1.70) | <0.001 |

NOTES: This table shows the results of multivariate logistic regression analyses that adjusted for the following covariates: sex, age, care needs level, and Charlson comorbidity index. The dependent variables were home-based end-of-life care utilization and medical institutional death. The exposure of interest was the home care facility type, with Group A as the reference category. Group A comprised users of enhanced Home Care Support Clinics/Hospitals (HCSCs) with beds, Group B comprised users of enhanced HCSCs without beds, Group C comprised users of HCSCs, and Group D comprised users of general clinics.

The distribution of medical and LTC service utilization per person-year across the groups is shown in table 3. The mean total number of care days used by the participants was similar across the four groups. However, there were significant inter-group differences when the number of days was categorized into inpatient care, outpatient care, and home care. The mean annual number of inpatient care days was highest in Group D (34.0 days), followed by Group B (33.0 days), Group C (29.6 days), and Group A (26.6 days). The mean annual number of outpatient care days was also highest in Group D (18.5 days), followed by Group A (10.1 days), Group C (9.8 days), and Group B (8.5 days). The mean annual number of home care days was highest in Group A (31.1 days), followed by Group C (27.2 days), Group B (24.9 days), and Group D (13.3 days). The mean annual inpatient care expenditure was highest in Group B (US$9,822.8) and lowest in Group A (US$7,661.7). The mean annual outpatient care expenditure was highest in Group D (US$1,109.3) and lowest in Group C (US$675.2). The mean annual home care expenditure was highest in Group A (US$6,122.2) and lowest in Group D (US$1,627.7). The mean annual prescription expenditure was highest in Group A (US$2,393.7) and lowest in Group D (US$1,722.2). The mean annual LTC expenditure was highest in Group A (US$30,252.7) and lowest in Group D (US$26,688.6).

**Table 3.** Medical and long-term care utilization and expenditure per person-year by home care facility type

|  | Group A | Group B | Group C | Group D | P value |
| --- | --- | --- | --- | --- | --- |
| Person-year | 4955.1 | 1703.5 | 13667.8 | 18564.3 |  |
| Mean [SD] | 2.0 [1.1] | 2.1 [1.1] | 2.2 [1.0] | 2.1 [1.0] |  |
| Rate per person-year |  |  |  |  |  |
| Care days |  |  |  |  |  |
| Inpatient care | 26.6 | 33.0 | 29.6 | 34.0 | <0.001 |
| Outpatient care | 10.1 | 8.5 | 9.8 | 18.5 | <0.001 |
| Home care | 31.1 | 24.9 | 27.2 | 13.3 | <0.001 |
| Care expenditure |  |  |  |  |  |
| Inpatient care | 7661.7 | 9822.8 | 8024.9 | 9382.9 | <0.001 |
| Outpatient care | 709.0 | 696.2 | 675.2 | 1109.3 | <0.001 |
| Home care | 6122.2 | 5172.5 | 4365.2 | 1627.7 | <0.001 |
| Prescription | 2393.7 | 2157.0 | 2151.9 | 1722.2 | <0.001 |
| Long-term care | 30252.7 | 29153.8 | 29457.1 | 26688.6 | <0.001 |

NOTES: Expenditures were converted from Japanese yen to US dollars using the 2017 purchasing power parity rate (US\$1 = 105.4). The values were calculated over the number of years each participant lived during the study period and are reported here as the annual rates per person-year of observation. Group A comprised users of enhanced Home Care Support Clinics/Hospitals (HCSCs) with beds, Group B comprised users of enhanced HCSCs without beds, Group C comprised users of HCSCs, and Group D comprised users of general clinics.

Table 4 shows the marginal means estimated from the GLMs evaluating the associations of home care facility type with medical and LTC service utilization. The number of inpatient care days was highest in Group D (75.0 days), followed by Group B (69.9 days), Group C (64.7 days), and Group A (54.3 days). The number of outpatient care days was also highest in Group D (40.2 days), followed by Group C (21.2 days), Group A (21.1 days), and Group B (17.1 days). In contrast, the number of home care days was highest in Group A (63.8 days), followed by Group C (57.8 days), Group B (51.0 days), and Group D (29.0 days). Inpatient care expenditure was highest in Group B (US$20,767.7), followed by Group D (US$20,413.7), Group C (US$17,606.3), and Group A (US$15,523.3). Outpatient care expenditure was highest in Group D (US$2,332.9), followed by Group A (US$1,522.8), Group C (US$1,500.6), and Group B (US$1,455.8). Home care expenditure was highest in Group A (US$12,747.4), followed by Group B (US$10,790.1), Group C (US$9,551.4), and Group D (US$3,440.9). Prescription expenditure was highest in Group A (US$5,183.1), followed by Group C (US$4,766.9), Group B (US$4,753.0), and Group D (US$3,715.1). LTC expenditure was highest in Group A (US$64,192.7), followed by Group C (US$64,147.1), Group B (US$62,003.3), and Group D (US$58,186.0). The results of the marginal means are also presented visually in figures 1 and 2.

**Table 4.** Comparison of medical and long-term care utilization and expenditure by home care facility type

|  | Care days |  |  | Care expenditure |  |  |  |  |
| --- | --- | --- | --- | --- | --- | --- | --- | --- |
|  | Inpatient | Outpatient | Home care | Inpatient | Outpatient | Home care | Prescription | Long-term care |
| Home care facility type |  |  |  |  |  |  |  |  |
| Group A | 54.3 | 21.1 | 63.8 | 15523.3 | 1522.8 | 12747.4 | 5183.1 | 64192.7 |
| Group B | 69.9 | 17.1 | 51.0 | 20767.7 | 1455.8 | 10790.1 | 4753.0 | 62003.3 |
| Group C | 64.7 | 21.2 | 57.8 | 17606.3 | 1500.6 | 9551.4 | 4766.9 | 64147.1 |
| Group D | 75.0 | 40.2 | 29.0 | 20413.7 | 2332.9 | 3440.9 | 3715.1 | 58186.0 |
| Sex |  |  |  |  |  |  |  |  |
| Men | 77.6 | 30.8 | 44.6 | 21639.5 | 2143.2 | 7867.3 | 4542.4 | 57792.6 |
| Women | 64.9 | 29.9 | 44.7 | 17681.4 | 1811.9 | 6885.3 | 4238.6 | 62216.6 |
| Age |  |  |  |  |  |  |  |  |
| 75–79 | 86.4 | 34.1 | 42.0 | 24272.6 | 2800.2 | 8101.9 | 5316.7 | 56446.0 |
| 80–84 | 76.2 | 32.2 | 43.3 | 21405.5 | 2271.6 | 7308.3 | 4789.7 | 60468.6 |
| 85–89 | 67.3 | 30.3 | 45.7 | 18527.4 | 1838.7 | 6957.2 | 4292.0 | 62223.8 |
| ≥90 | 58.3 | 26.9 | 45.6 | 15569.4 | 1326.0 | 6805.5 | 3598.5 | 62691.4 |
| Care needs levels |  |  |  |  |  |  |  |  |
| Level 3 | 64.2 | 30.7 | 41.0 | 18907.6 | 2161.7 | 6706.5 | 4518.9 | 53625.4 |
| Level 4 | 68.7 | 31.5 | 43.4 | 18701.5 | 2023.9 | 6622.3 | 4238.5 | 60912.2 |
| Level 5 | 71.6 | 28.0 | 49.2 | 18829.2 | 1516.9 | 8029.8 | 4202.8 | 68709.6 |
| Charlson comorbidity index |  |  |  |  |  |  |  |  |
| 0–2 | 49.2 | 28.5 | 41.3 | 12987.0 | 1351.7 | 5854.5 | 3405.7 | 62673.7 |
| 3–4 | 65.0 | 29.6 | 44.4 | 17513.9 | 1648.3 | 6610.3 | 4109.3 | 62362.6 |
| ≥5 | 81.0 | 31.5 | 46.8 | 22791.9 | 2418.9 | 8247.3 | 5001.0 | 59337.2 |
| Death |  |  |  |  |  |  |  |  |
| No | 42.8 | 25.7 | 40.3 | 12031.3 | 1607.1 | 6180.9 | 3766.4 | 54020.8 |
| Yes | 124.9 | 40.8 | 53.4 | 32308.4 | 2684.7 | 8988.1 | 5831.8 | 81395.7 |

NOTES: Expenditures were converted from Japanese yen to US dollars using the 2017 purchasing power parity rate (US\$1 = 105.4). The table shows the results (marginal means) of generalized linear models assuming a negative binomial distribution for care days and a gamma distribution for care expenditure.
Analyzed, dependent variables include: inpatient care days, outpatient care days, home care days, inpatient care expenditures, outpatient care expenditures, home care expenditures, prescription expenditures, and long-term care expenditures. The exposure of interest was the homecare facility type. The models adjusted for the following covariates: sex, age, care needs level, Charlson comorbidity index, death, and the number of years the participants lived during the study period. The marginal means of the dependent variables were calculated by substituting the means of the estimates into the generalized linear regression models. Group A comprised users of enhanced Home Care Support Clinics/Hospitals (HCSCs) with beds, Group B comprised users of enhanced HCSCs without beds, Group C comprised users of HCSCs, and Group D comprised users of general clinics.

**Image1: Figure1.**
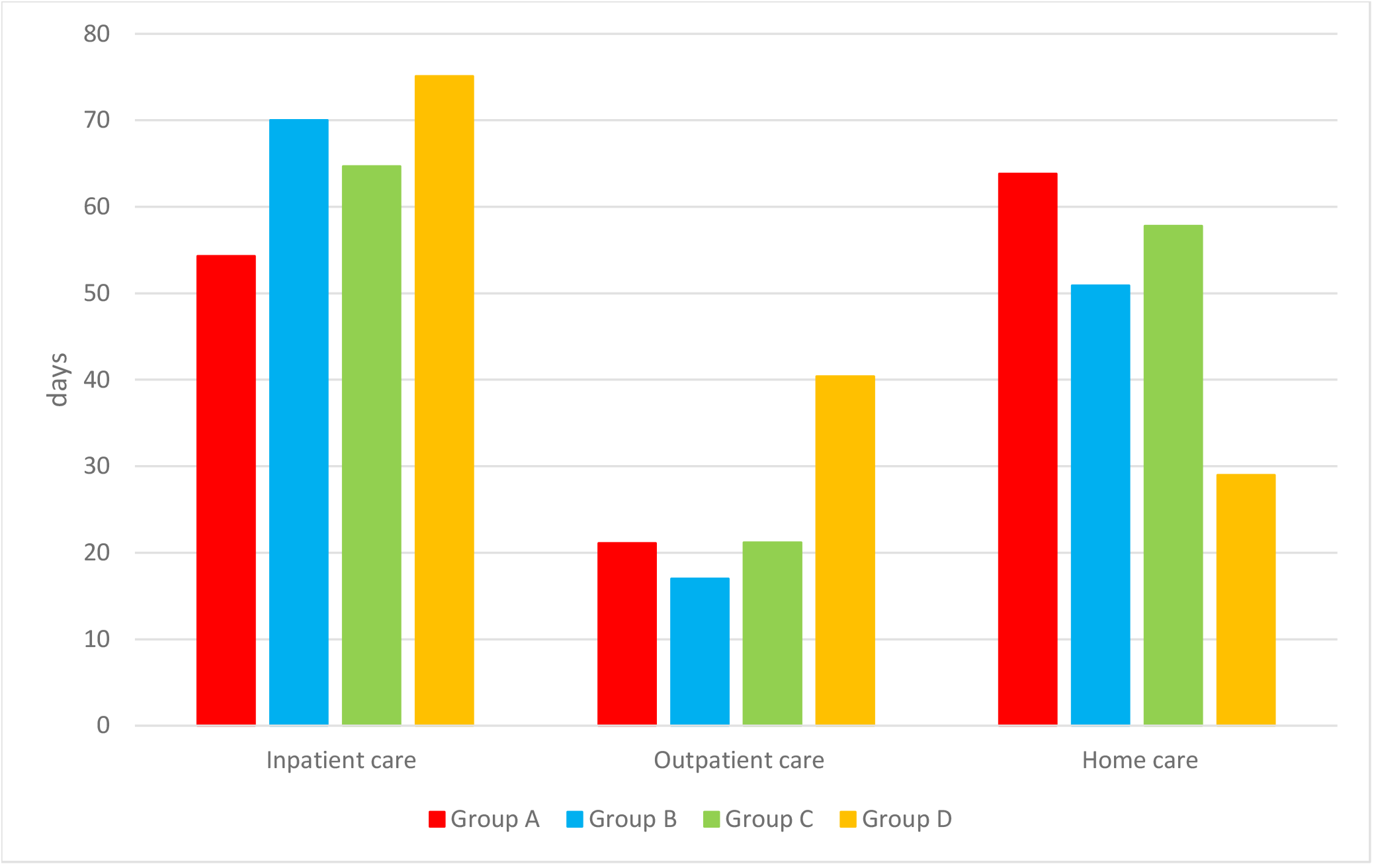
The title is “Comparison of medical utilization using marginal means by home care facility type”. This shows the care days by home care facility, which is the result of Table 4.

**Image2: Figure2.**
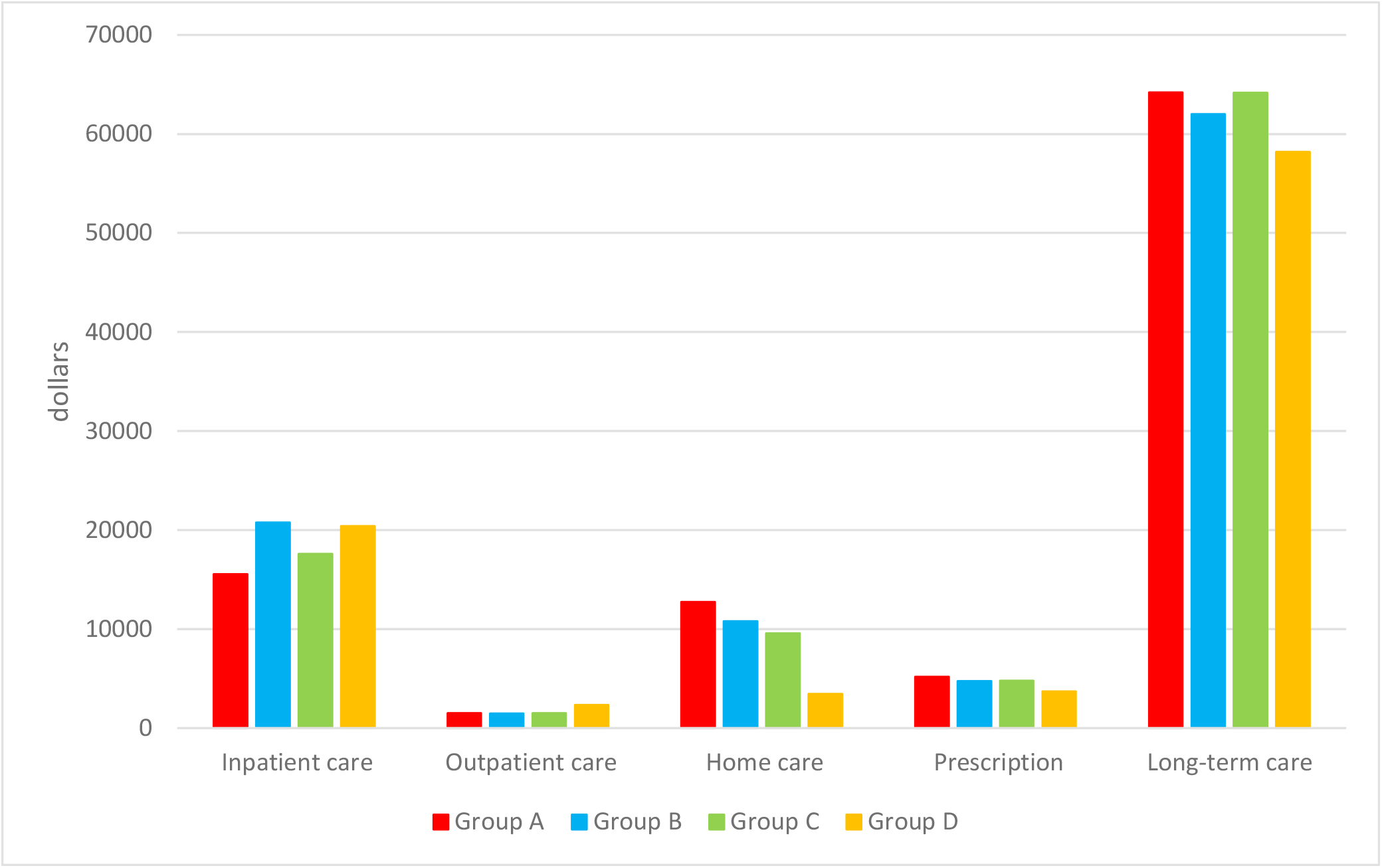
The title is “Comparison of medical and long-term care expenditures using marginal means by home care facility type”. This shows the care expenditures by home care facility, which is the result of Table 4.

## DISCUSSION

In this retrospective study of older adult home care service users with high care needs levels residing in Fukuoka Prefecture, we compared the utilization of medical and LTC services among patients treated by four types of home care facilities. Participants who used enhanced HCSCs with beds had the highest number of home care days and the lowest number of inpatient care days per person-year. In contrast, participants who used general clinics had the lowest number of home care days and the highest number of inpatient and outpatient care days per person-year. These results corroborate previous findings showering that the integration of home care into community care by specialized clinics is effective in reducing hospitalization duration among older adults.[20, 34]

A novel finding of the present study is that participants who used home care services from enhanced HCSCs with beds were the most likely to receive end-of-life care at home and the least likely to die in a medical institution. Specifically, the odds of institutional death were 2.32 times higher for participants who used general clinics than for those who used enhanced HCSCs with beds, indicating that treatment by the latter type of facility is associated with a reduction in medical institutional deaths. The Ministry of Health, Labour and Welfare of Japan reported that the national medical institutional death rate in 2016 was 75.8%,[35] which was substantially higher than the corresponding rate observed among the patients in our study (36.6%) and even among the general clinic user group alone (44.3%). This suggests that older adults who use home care services are more likely to die outside of medical institutions, in places such as their homes or an LTC facility. Promoting the use of home care could, therefore, help to reduce the medical institutional death rate, as has also been demonstrated in a previous study.[36] Similarly, Sadamura and Babazono examined the correlation between LTC resources and place of death, finding that collaborations among clinics to provide home-based medical and LTC services reduced medical institutional deaths.[37] This form of integrated care may be effective in providing home-based end-of-life care for older adults.

It is noteworthy that participants who used enhanced HCSCs with beds had a substantially lower number of inpatient care days and expenditures but a higher number of home care days and expenditures, compared with participants who used enhanced HCSCs without beds. This suggests that the presence of beds in enhanced HCSCs affected the use of care, despite HCSCs with and without beds being otherwise functionally identical. This suggests that enhanced HCSCs with beds are more likely identified as hospitals rather than privately owned clinics, that are focused on providing home care despite having an abundance of health care resoures for providing inpatient care. Another notable result is that conventional HCSCs appeared to be more effective in providing home care, compared with enhanced HCSCs without beds in terms of the metrics of inpatient care, home care days, and home care expenditures. These findings indicate a need to review the current insurance system, which reimburses enhanced HCSCs without beds at a higher rate for the same services, compared with conventional HCSCs.[15] In our study population, the users of enhanced HCSCs with beds had a higher utilization of home-based end-of-life care and a lower utilization of inpatient care, despite having relatively high care needs levels. Among the home care facility types, those that are well equipped to manage sudden changes in their patients’ conditions may be able to provide high-quality home-based daily living support and reduce the length of hospitalization. A point of concern is that there was no significant difference in the number of inpatient care days between general clinics and enhanced HCSCs without beds. The prerequisites for receiving enhanced HCSC status may be met by a home care facility through collaboration with other clinics.[15] Therefore, it is possible that some of the enhanced HCSCs without beds were not individual facilities but instead comprised two or more clinics collaborating to satisfy the relevant criteria. Consequently, decisions on treatment strategies (such as hospitalization) at these facilities may involve doctors working at several facilities rather than only one facility. Bynum et al. examined the effects of different types of primary care on hospitalizations in a continuing care retirement community, finding that individuals with 24-hour primary care availability from physicians providing care only at that site had significantly fewer hospitalizations and emergency department visits than did those who were served by external non-site-specific doctors.[34] This suggests that the decentralization of healthcare provision may be reduced when one facility is in charge of providing primary care for older adults living in a community, which can improve the quality of care and reduce the number of hospitalizations and overall healthcare utilization rates. On the basis of our findings and those from previous studies, we propose that the home care delivery system in Japan to encourage the development of “enhanced” status to clinics with beds. Under Japan’s current medical fee schedule, there are no differences in home care fees between enhanced facilities operating independently and those working in collaboration with other facilities. To expand the role of enhanced HCSCs, when examining the differences among HCSCs, it is necessary to consider whether the facility works in collaboration with other clinics in addition to considering whether the facility has beds.

This study has several limitations. First, the study was conducted using data only on Fukuoka Prefecture residents, which limits the generalizability of our findings.[38] This prefecture has a relatively high number of hospital beds and relatively high medical expenditures per person, and this study’s results therefore may be overestimated. Second, our data did not include detailed information about living conditions reflecting the participants’ family structure or characteristics of living, which may influence the choice of a home care facility. Third, although the statistical analyses incorporated characteristics such as sex, age, care needs level, and CCI, the specific diseases of each participant were not taken into consideration. Fourth, no clinical data (e.g., disease progression or laboratory test results) for individual participants were included because this study focused on the types of healthcare facilities providing home care. Moreover, the issue related to the possibility of participants who moved from home to a long-term care facility such as a nursing home during the follow-up period was not addressed. Finally, we used care needs level as a covariate, but we were unable to account for any changes in this level over the study period. Nevertheless, considering the care needs level and CCI at the start of the study provided insight into the participants’ baseline disease severity. In this study, we showed the difference of home care system in Japan’s CICS on the use of medical and LTC services, the use of home-based end-of-life care, and the place of death among older community-dwelling adults. We also confirmed the important role of enhanced HCSCs with beds in providing home care services. Currently, there are 7,629 clinics with beds in Japan, and approximately 100,000 beds are available. Of these beds, 46% are used for emergency care and 37% provide transitional care for hospital-discharged patients before they are transferred to home or to an LTC facility.[39] Approximately 60% of the patients occupying the available beds are aged 75 years and older.[39] To optimize the provision of home care through a community-based care system, it is necessary to consider functional changes in clinics with beds. The promotion of integrated community care is regarded as a viable solution for aging societies in many countries. To support the increasing need for community care, the World Health Organization published *Integrated Care for Older People: Guidelines on Community-level Interventions to Manage Declines in Intrinsic Capacity* in 2017.[40] These guidelines, which emphasize the need for comprehensive community-based strategies and primary care-level interventions to prevent diminishing capacity, are consistent with our study’s conclusion that HCSCs with beds play an integral role in Japan, as the main healthcare facilities providing home care. As part of its national policy, Japan is considering a further expansion of the CICS. This expansion would involve the construction of a large system by coordinating the resources of acute care hospitals, rehabilitation hospitals, LTC facilities, clinics with beds, primary healthcare clinics, and comprehensive support centers within each region of the country. Consequently, there would be a need to clarify each facility’s role in this expanded system. Our study provides useful information for further investigations of home care for older adults as part of community-based integrated care.

## Data Availability

No additional data are available.

## Acknowledgements

The authors thank the Fukuoka Prefecture Association of Latter-Stage Elderly Healthcare for provision of the healthcare claims database. We also thank Jennifer Barrett, PhD, from Edanz Group (https://en-author-services.edanzgroup.com/ac) for editing a draft of this manuscript.

## Contributors

SK lead the study design, conducted the literature search, extracted and analyzed the data, and wrote the manuscript. AB contributed to the study design, analysis and manuscript revision. AJ, YL and LN contributed to the study design and analysis.

## Patient consent for publication

Not required.

## Ethics approval

This study was approved by the Institutional Review Board of Kyushu University (Clinical Bioethics Committee of the Graduate School of Healthcare Sciences, Kyushu University). (Approval No. 20209).

## Provenance and peer review

Not commissioned; externally peer reviewed.

## Data sharing statement

No additional data are available.

## Open access

